# Men’s preconception health and the social determinants of health: What are we missing?

**DOI:** 10.1101/2022.05.24.22275546

**Authors:** Adaobi Anakwe, Hong Xian, Rhonda BeLue, Pamela Xaverius

**Affiliations:** Department of Health Sciences, School of Health Professions, University of Missouri Columbia, Missouri, USA, 65211; Department of Epidemiology and Biostatistics, College for Public Health and Social Justice, Saint Louis University, USA, 63104; College for Health, Community and Policy, University of Texas San Antonio, Texas, 78249; University of Health Sciences and Pharmacy, St. Louis, Missouri, 63110

**Author notes:** **Corresponding author** Adaobi Anakwe, Rm 315 Clark Hall, University of Missouri, Columbia MO, 65211. **Funding information** Funding for this study was received through the Society for Family Planning Research Fund (Grant Number: SFPRF14-ES2). **Author contributions** AA: Conceptualized and designed the study, analyzed data, and drafted the initial manuscript RB: Contributed to study conceptualization and design, analysis, and critically manuscript revisions HX: Contributed to data analysis and interpretation PX: Contributed to study design and critically revised the manuscript for intellectual content. **Data availability** The data used in this study are publicly available through the National Center for Health Statistics website. The datasets generated and/or analyzed during the current study are available in the Harvard Dataverse repository, https://doi.org/10.7910/DVN/4X4IEG. **Contributions to the field** This study used pooled data from the 2011-2019 National Survey of Family Growth to examine the relationship between men’s preconception health and the social determinants of health. It specifically highlighted the need to consider preconception health as a latent construct by examining the constellation of co-occurring risk factors that characterize men’s preconception health, and as they are predicted by the social determinants of health. To our knowledge, this is the first paper that applies this method to measure and examine the effect of the SDOHs on the men’s preconception health at the population level. Findings identified three latent classes of men’s preconception health – low risk, substance users, and sexual risk-takers - and demonstrated that health access, socioeconomic status, and sociocultural contexts, but not residential context were significant predictors of class membership. Our study specifically highlights that higher socioeconomic status does not protect men from poor preconception health which underscores the need for more in-depth examinations of the relationship between social class, masculinity, and men’s preconception health. It further suggests that other underlying factors that are nested in men’s experiences of their social contexts influence men’s preconception health.

**Keywords:** preconception health, latent class analysis, social determinants, quantitative, men

## Abstract

**Background:** Life course perspectives suggest that optimizing men’s health before conception is requisite to equitably improve population health, an area of increasing public health focus. Although scholarship on the social determinants of health (SDOH) suggests that men’s health and health behaviors do not occur in a vacuum, preconception health studies have not explicitly examined how these factors influence men’s preconception health.

**Objective:** To identify latent classes of men’s preconception health and the role of the SDOHs in predicting class membership.

**Methods:** Pooled data from the 2011-2019 male file of the National Survey of Family Growth were analyzed (n= 10,223). Latent class analysis was used to identify distinct classes of men’s preconception health. Eight manifest variables were used to fit latent class models. A classify-analyze approach was subsequently used to create a preconception health phenotype (*PhP)* outcome variable. SDOHs (exposure variable) were assessed in four domains (residential context, health access, socioeconomic status, and sociocultural context) to predict class membership. Survey weighted multinomial regression models were fitted to examine the association between the exposure and the outcome.

**Results:** Three unique *PhP*s were identified (lowest risk (69%), substance users (22.9%), and sexual risk-takers (8.1%) (SRT)) from the LCA model. Health access, socioeconomic status, and sociocultural contexts were significant predictors of class membership but not residential context. Sexual risk takers were more likely to be uninsured (aOR: 1.25, 95% CI 1.02, 1.52), college-educated (aOR: 1.94 95% CI: 1.34, 2.79), and non-Hispanic Black (aOR: 1.99 95% CI: 1.55, 2.54) while substance users were more likely to have unstable employment (aOR: 1.23 95% CI:1.04, 1.45) and have a high school degree or higher (aOR 1.48 95% CI: 1.15, 1.90) than men in the lowest risk category.

**Conclusion:** Findings suggest that social determinants may impact men’s preconception health in ways that are not conventionally understood and raises important questions about how preconception health interventions should be created, tailored, and/or retooled. Specifically, studies that examine the sociocultural and political contexts underpinning the relationship between social class, masculinity, and men’s preconception health are needed to provide nuanced insights on factors that shape these outcomes.

## BACKGROUND

Preconception health, defined as the health of women and men from pubarche to when they can have a child [1], is critical to improving population health across the life course. However, most preconception health research disproportionately focuses on women with the greatest progress towards understanding the salience of men’s preconception health made only in the past 10 years. These studies, which often focus on men’s biological contributions to pregnancy, suggest that men’s poor preconception health can impair fertility, and reduce semen quality and quantity[2-7]. Less is known about the social factors that contribute to men’s preconception health or how these factors impact the aggregation of preconception health indicators at the population level. In 2006, the CDC recommended ten surveillance indicators that could optimize the health of men preconceptionally. These indicators include: making a reproductive plan, preventing sexually transmitted diseases, quitting substance use, avoiding exposure to toxic substances, preventing infertility maintaining a healthy weight, knowing family history, seeking help for violence, staying mentally healthy, and supporting partners [8].

Despite the awareness of these factors, optimizing preconception health is constrained by a host of factors including a) an inconsistent definition of the preconception period b) difficulties with making and sustaining behavior change without a clear understanding of the interactive effects of contextual factors c) the persistently high prevalence of unintended pregnancies and d) persistent disparities in access to and utilization of health care resources [9-11]. In addition to these factors, men face other unique challenges to optimizing their health preconceptionally. For instance, compared to women, men are more likely to engage in risky behaviors and less likely to modify these behaviors over time, less aware of the health behavior changes they need to make or how these behaviors can impact their offspring and are less inclined to seek health care [12-15].

Men’s health and health behaviors do not occur in a vacuum. Research has increasingly acknowledged the role of the social determinants of health - i.e., where people are born, live, play, learn, work, pray, and age - in creating and shaping health trajectories across the life course [16]. In 2018, about 9% of men between the ages of 18 to 64 lived in poverty, 10.6% had less than high school education, and 3.4% of men 20 years and over were unemployed [17-19]. Of these, racial and ethnic minorities accounted for the greater proportion of those at lower socioeconomic status [poverty rate: non-Hispanic Black (20.8%), Hispanics (17.6%), and foreign-born populations (13.8%)]. Among non-Hispanic Black and Hispanic males, those with less than a high school degree accounted for 11.7% and 29.3% respectively [17, 18]. These minority populations often report poorer health than their non-Hispanic White counterparts. Social factors intertwine with biological factors across the life course to shape health outcomes, which can adversely impact men’s preconception health [20, 21]. Given the stark social disparities that underlie men’s health, there is a need to examine how men’s social environment influences their preconception health status with attention to the neighborhood, health access, and socioeconomic and social/community factors.

The objective of this study, therefore, was to utilize data from the National Survey of Family Growth (NSFG) to a) characterize latent clustering of men’s preconception health at the population level and b) examine the extent to which the social determinants of health - residential context, health access, socioeconomic status and social and community factors influence these clustering. Preconception risk factors such as risky sexual behaviors and substance use or chronic disease and poor nutrition often co-occur and are intertwined with social factors like socioeconomic status and racial/ethnic background [22, 23]. Therefore, this study hypothesized that a) latent classes of men’s preconception health will be observed and b) that men’s preconception latent class membership will be predicted by the social determinants of health.

## MATERIALS AND METHODS

### Study Participants and Data Source

Pooled data from the National Survey for Family Growth (NSFG) from 2011 to 2019 was used. The NSFG is a multi-stage, stratified, probability sample of the non-institutionalized U.S. with in-person interviews conducted continuously at 2-year intervals. Details on data collection methods are reported elsewhere [24]. Data were collected from both males and females ages 15-44 years (extended to 49 years in the 2015-2019 cycles) with data collection among men only beginning in 2002. Men included in this study were sexually experienced with a female, fecund, and had at least one fecund partner [25, 26]. Those who also reported a current pregnant partner were excluded. These criteria were selected to identify those who at the time of data collection, were at risk of experiencing a pregnancy with their partner. Data were weighted to account for the complex survey and provide estimates that are generalizable to the U.S male population.

### Measures of Men’s Preconception Health Status

Men’s preconception health was measured with eight lifestyle variables – the number of sexual partners, sexual risk-taking behavior, condom use consistency, general health status, alcohol, and drug use, exposure to sexually transmitted infections (STI), and body mass index. These variables were further categorized into 5 domains - sexual behavior and awareness, medical history, substance use history, infectious disease status, and healthy weight. Only these five domains were assessed because of the availability of these variables in the NSFG data. These manifest variables were dichotomized following sensitivity analysis which showed no significant differences between the categories by sociodemographic characteristics. A detailed description of these variables is provided elsewhere (Authors – BLINDED FOR REVIEW).

#### Sexual behavior and awareness

Men’s sexual risk behavior was measured using three variables that assessed the number of sexual partners, condom use, and STI/HIV risk-taking behavior. The first variable assessed the number of female partners a man had with the question *“number of female partners in the last 12 months”* and was recoded as 1= none or one female partner, and 2=more than one female partner [26]. The question, *“in the last 12 months, how often did you use a condom with your partner or partners”* was used to measure condom use. These responses were recoded to measure condom use consistency where 1= used consistently and 2= inconsistent use to no use [25]. Sexually transmitted infection and HIV risk-taking behaviors were measured using five questions that asked whether a man a) had sex with a female intravenous drug user, b) gave money or drugs to a female for sex, c) took money or drugs from a female for sex, d) had sex with an HIV-positive female, and e) had any other sexual experience with another man with a binary (yes/no) response option. The first four questions inquired about past 12-months exposure whereas the fifth question inquired about lifetime exposure. These variables were used to create a single STI/HIV risk dummy variable (1=No, 2=Yes) [26].

#### Medical History

Men’s general health status was used as a proxy measurement for medical history and was measured using the question “In general, how is your health?” with responses on a five-point scale ranging from excellent to poor. For analysis, this variable was recoded to a dummy variable 1= excellent to good, 2= fair to poor. Since only the 2015-2019 NSFG files measured high blood pressure and medication use among males, these variables were not used for this analysis. The general health status measure is the most commonly available measure of perceived overall health in national surveys and previously showed fair to good test-retest reliability in a population study [27].

#### Substance Use History

Alcohol use was measured using three questions that assessed the frequency and quantity of alcohol use. These variables were recoded as “no drinking” if no alcohol use was reported in the past 12 months and 30 days, “low risk” if alcohol use was reported in the past 30 days but not at binge levels, “medium risk” if they binge drank less than 5 times in the past 30 days and “high risk” if they drank 5 or more times in the past 30 days [28]. For analytical purposes, these risk categories were further recategorized into two risk groups: 1 = no to low-risk drinking and 2 = medium to high-risk drinking. Six questions on drug use inquired about marijuana, cocaine, crack, methamphetamine, and injection drug use in the past 12 months. These variables were recoded into a single “any drug use” category with a binary (yes/no) response. Since only the 2015-2019 NSFG questionnaire collected responses on smoking status, substance use history was measured using the respondent’s alcohol and drug use history only.

#### Infectious Disease Status

Five questions were used to measure the presence or absence of sexually transmitted infections. These questions asked whether the respondent was told that they had gonorrhea, chlamydia, herpes, genital warts, and/or syphilis in the last 12 months. Responses to these questions were used to create a single STI status dummy variable with a binary (yes/no) response.

#### Body Mass Index

Body mass index is the most commonly used marker to ascertain healthy weight status [29]. Body mass index was collected as a continuous variable and was categorized into 4 distinct groups – underweight (less than 18.0), Normal weight (19 to <25), overweight (26 to <29), and obese (30 or higher)[30]. These were recategorized further into a dichotomous variable “under to normal weight” and “overweight to obese.”

### Social determinants of men’s preconception health (predictor variables)

Residential Context was measured using participants’ place of residence with a 3-item response scale that was determined by the Metropolitan Statistical Area (1=Principal city of MSA, 2=Other MSA, and 3=not MSA) that we renamed as urban, suburban, and rural residence respectively.

Health access was measured with the question “In the last 12 months was there any time that you did not have any health insurance or coverage?” with a binary (yes/no) response. Health care utilization was measured with two questions that assessed the type of doctor’s visit (1=routine physical exam, 2=a physical exam for sports or work, 3=a doctor visit when you were sick or hurt, 4=did not have any visits to a doctor) and whether the doctor’s visit was for reproductive health care. These variables were recoded as “reproductive wellness visit” if the participants went to the doctor and received reproductive health information, screening, exam, or treatment; “no reproductive wellness visit” if they went to the doctor for reasons other than for reproductive health and “no visit” if they did not have any doctor’s visit in the past 12 months

Socioeconomic factors were measured in three domains - employment consistency, poverty level, and educational attainment. Employment consistency was examined for the past 12-months and was categorized as “unemployed” if the respondent was not employed in the last 12 months, “unstable employment” if the respondent was employed for less than 12 months, and “stable employment” if the respondent was employed for 12 months. The federal poverty-to-income ratio (PIR) grouped into two subcategories - at or below 100% of family poverty level versus above 100% of the family poverty level – was used to determine poverty level [31]. Educational attainment was measured on a four-point scale - less than high school, high school, some college, and college degree or more.

Racial/ethnic belonging and immigration status were used as a proxy measure for social and cultural contexts. Immigration status is fast becoming an acknowledged social determinant of health and was a dichotomized variable (i.e., immigrant versus non-immigrant) utilized in this study to measure the underlying cultural and normative beliefs and practices around preconception health. Race/ethnicity was a four-level categorical variable - non-Hispanic White, non-Hispanic Black, Hispanic and Other.

### Covariates

Covariates were sociodemographic characteristics that were independently associated with the exposure, outcome, or both variables and relevant to the literature. These included union type (married, separated/widowed/divorced, and never married), age, number of children the ever fathered, and age of sexual debut.

### Statistical analysis

Descriptive statistics including frequencies and bivariate analysis were used to describe the preconception health indicators and sociodemographic factors.

A latent class analytical approach was used to characterize men’s preconception health. Latent class analysis (LCA) is a statistical technique used to observe unobserved phenomena and identify subgroups of individuals in a set of two or more mutually exclusive and exhaustive latent classes based on multiple observed variables [32]. This method was used because of the heterogeneity in preconception health behaviors, interrelatedness between these risk factors, and the utility of this method to identify unique dimensions of preconception health from these variables. An iterative maximum likelihood method was used to specify two parameters: (1) the prevalence of latent classes (Gamma parameters) and (2) the item response probability representing the probability of endorsing a particular item within each class.

The LCA model was fitted starting with the 2-class model with a one-unit increment in the number of classes (3-, 4-, and 5-class models). The model of best fit was determined based on the Bayesian Information Criterion (BIC), entropy, mean posterior probabilities and class size [32], the interpretability of the solution, parsimony of the model, and relevance to the literature. The smaller BIC value indicates a better model fit. Entropy denotes how accurately a model defines a class with values closer to 1 considered ideal [33]. Mean posterior probabilities are presented in a matrix with probabilities of membership in each class on the diagonal. Diagonals close to 1 (0.8 and above) and off-diagonals close to zero are considered more reliable models. Analyses were completed using the PROC LCA procedure in SAS 9.4 [34]. The combined 8-year weight for the 2011-2019 period was applied to all analyses.

Using the posterior probabilities derived from the latent class models, a preconception health phenotype (*PhP*) variable was created, by applying a classify-analyze approach [33] and was subsequently used to fit multinomial logistic regression models to determine the relationship between *PhP* and the social determinants of health. Although “phenotype” is a biological term that refers to the physical expression of genes, it was applied to this study to denote the set of observable characteristics that distinguished members within a specific latent class from those in other latent classes. Regression models estimated odds ratios with corresponding 95% CI. Independent crude and adjusted regression models were fitted for each social determinant. Statistical significance was placed at a p-value <.05 and 95% CIs not overlapping 1.0.

## RESULTS

### Descriptive statistics

A total of 10,223 men were included in the final sample and had an average age of 30.7(SE=0.13) years. Most men were non-Hispanic White (57.4%), never married (55.4%), had some college education (32.0%) with stable employment (72.4%). The majority also had health insurance (71.1%) and reported good to excellent health (93.9%). In line with the objectives of this study, the pooled eight-year sample was used to identify the weighted frequencies of positive endorsement of the preconception risk manifest variables. Overall, more than 80% of men reported inconsistent to no condom use, 61.9% reported being overweight or obese and 18.6% reported having two or more sexual partners. About a third (32.2%) used drugs and 41.7% used alcohol at medium to high-risk levels (Table 1).

**Table 1:**
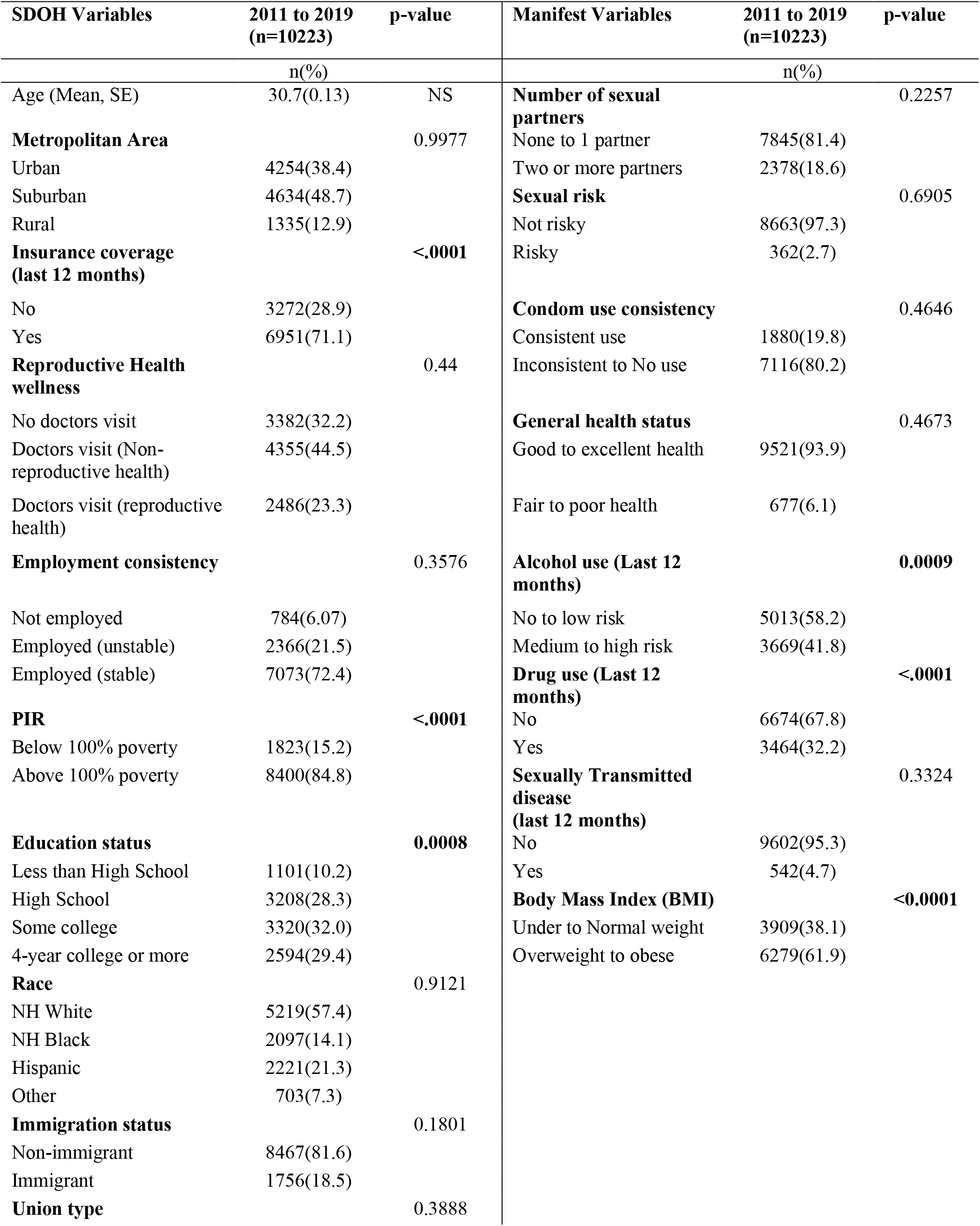
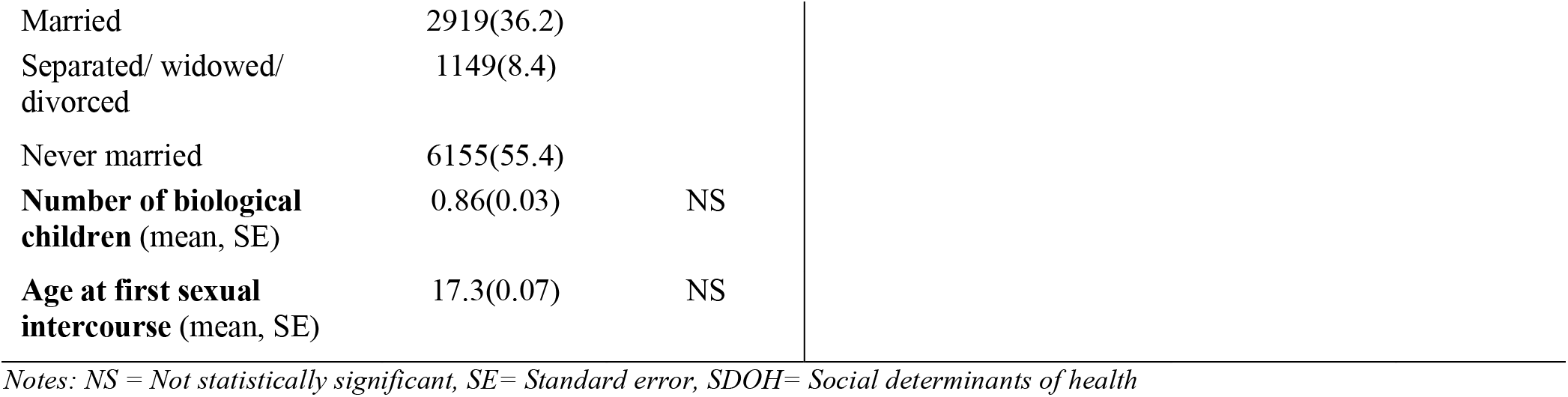
Weighted descriptive statistics of National Survey of Family Growth male sample (2011-2019)

| SDOH Variables | 2011 to 2019<br>(n=10223) | p-value | Manifest Variables | 2011 to 2019<br>(n=10223) | p-value |
| --- | --- | --- | --- | --- | --- |
|  | n(%) |  |  | n(%) |  |
| Age (Mean, SE) | 30.7(0.13) | NS | **Number of sexual partners** |  | 0.2257 |
| **Metropolitan Area** |  | 0.9977 | None to 1 partner | 7845(81.4) |  |
| Urban | 4254(38.4) |  | Two or more partners | 2378(18.6) |  |
| Suburban | 4634(48.7) |  | **Sexual risk** |  | 0.6905 |
| Rural | 1335(12.9) |  | Not risky | 8663(97.3) |  |
| **Insurance coverage (last 12 months)** |  | <.0001 | Risky | 362(2.7) |  |
| No | 3272(28.9) |  | **Condom use consistency** |  | 0.4646 |
| Yes | 6951(71.1) |  | Consistent use | 1880(19.8) |  |
| **Reproductive Health wellness** |  | 0.44 | Inconsistent to No use | 7116(80.2) |  |
| No doctors visit | 3382(32.2) |  | **General health status** |  | 0.4673 |
| Doctors visit (Non-reproductive health) | 4355(44.5) |  | Good to excellent health | 9521(93.9) |  |
| Doctors visit (reproductive health) | 2486(23.3) |  | Fair to poor health | 677(6.1) |  |
| **Employment consistency** |  | 0.3576 | **Alcohol use (Last 12 months)** |  | **0.0009** |
| Not employed | 784(6.07) |  | No to low risk | 5013(58.2) |  |
| Employed (unstable) | 2366(21.5) |  | Medium to high risk | 3669(41.8) |  |
| Employed (stable) | 7073(72.4) |  | **Drug use (Last 12 months)** |  | <.0001 |
| **PIR** |  | <.0001 | No | 6674(67.8) |  |
| Below 100% poverty | 1823(15.2) |  | Yes | 3464(32.2) |  |
| Above 100% poverty | 8400(84.8) |  | **Sexually Transmitted disease (last 12 months)** |  | 0.3324 |
| **Education status** |  | **0.0008** | No | 9602(95.3) |  |
| Less than High School | 1101(10.2) |  | Yes | 542(4.7) |  |
| High School | 3208(28.3) |  | **Body Mass Index (BMI)** |  | <**0.0001** |
| Some college | 3320(32.0) |  | Under to Normal weight | 3909(38.1) |  |
| 4-year college or more | 2594(29.4) |  | Overweight to obese | 6279(61.9) |  |
| **Race** |  | 0.9121 |  |  |  |
| NH White | 5219(57.4) |  |  |  |  |
| NH Black | 2097(14.1) |  |  |  |  |
| Hispanic | 2221(21.3) |  |  |  |  |
| Other | 703(7.3) |  |  |  |  |
| **Immigration status** |  | 0.1801 |  |  |  |
| Non-immigrant | 8467(81.6) |  |  |  |  |
| Immigrant | 1756(18.5) |  |  |  |  |
| **Union type** |  | 0.3888 |  |  |  |

|  |  |  |
| --- | --- | --- |
| Married | 2919(36.2) |  |
| Separated/ widowed/<br>divorced | 1149(8.4) |  |
| Never married | 6155(55.4) |  |
| **Number of biological**<br>**children** (mean, SE) | 0.86(0.03) | NS |
| **Age at first sexual**<br>**intercourse** (mean, SE) | 17.3(0.07) | NS |

*Notes: NS = Not statistically significant, SE= Standard error, SDOH= Social determinants of health*

### Latent classes of men’s preconception health

The 3-class model provided the best fit across the eight preconception health manifest variables given 50 random start values, highest entropy values, and separation into distinct classes based on posterior probabilities of class membership (probability ranging from 0.84 to 0.97 within the class). Since a classify-analyze technique was applied to create a preconception health variable, it was important that the mean posterior probabilities for class membership be high to reduce the effect of random error (Supplementary material eTable 1a and 1b).

Table 2 represents the latent class probabilities (gamma estimates) i.e., the proportion of men expected to belong in each latent class, and the item response probabilities i.e., the proportion of men who positively endorsed these preconception health indicators within each class. Most men (69%) belonged to Class 1 “lowest risk group.” This class was characterized by a high endorsement probability of inconsistent /no condom use (82%) and overweight/obese (65.3%); all other prevalence estimates for each manifest variable were low. Class 2 “sexual risk-takers” had the lowest prevalence of latent classes (8.1%) and was characterized by high endorsement probability for multiple sexual partners (94.2%), inconsistent/no condom use (68.2%), medium/high alcohol use (54.1%) and overweight/obese (63.8%). Approximately 23% of men belonged to Class 3 “substance users” and were characterized by high endorsement probability for inconsistent/no condom use (79.6%), medium/high-risk alcohol use (66.9%), drug use (98.8%), and overweight/obese (50.8%). Item response probabilities >0.50 were selected to characterize and facilitate interpretation of class membership. Because men across all three classes had a high probability of endorsing inconsistent condom use and overweight/obese, each additional class was subsequently labeled based on an additional preconception risk factor that characterized class membership.

**Table 2:**
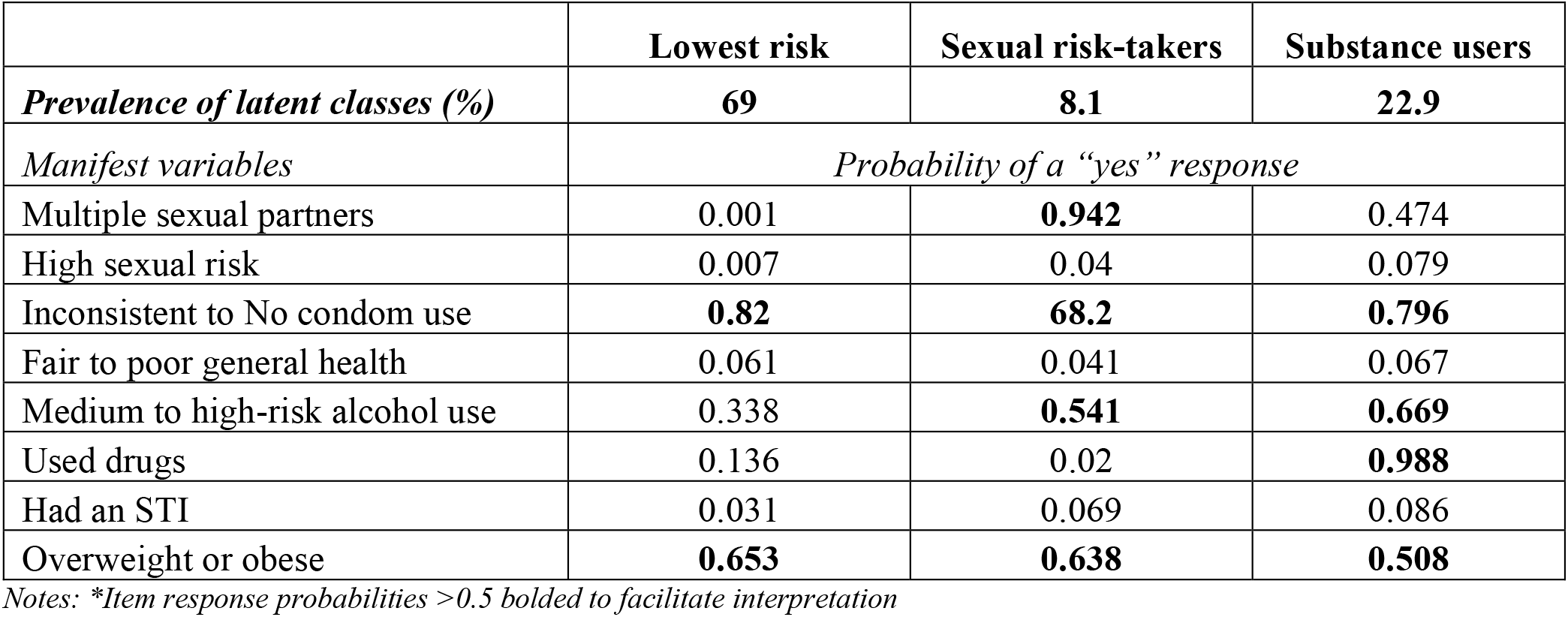
Latent classes of preconception health from the 3-class model

### Latent class structure and the social determinants of health

A description of the study sample, as they differed by the social determinants of health within their distinct preconception health groups, is provided in Table 3. More men in the “lowest risk” category lived in suburban areas (51%), whereas more men in the “sexual risk-takers” group lived in urban centers (44.3%). More men in the “substance users” category lived in suburban areas (47.3%). Across the *PhP* categories, most men were insured and had non-reproductive health visits to the doctor. Most men had stable employment, however, 30.7% of men in the “substance users” category had unstable employment, and 22.2% of “sexual risk-takers” also had unstable employment. Most men in the “lowest risk” category had a college degree or more (32.2%), whereas most substance users (36%) and sexual risk-takers (36%) had finished some college. Across all *PhP*s, most non-Hispanic White men belonged to the “sexual risk-takers” category while the majority of non-Hispanic Black men belonged to the “substance users” category. There was an even distribution of Hispanic men in the “lowest risk” and “substance users” categories. Immigrants were more frequently in the “lowest risk” group compared to non-immigrants.

**Table 3.** Weighted distributions of social determinants of health by men’s preconception health phenotypes.

| Variables | Preconception health latent classes |  |  | P-value |
| --- | --- | --- | --- | --- |
|  | Lowest risk<br>(n=6,652) | Sexual risk takers<br>(n=2,479) | Substance users<br>(n=1092) |  |
|  | n(%) | n(%) | n(%) |  |
| **Metropolitan area** |  |  |  | **<.0001** |
| Urban | 2644 (36.4) | 1170 (44.3) | 440 (38.9) |  |
| Sub-urban | 3155 (51.0) | 993 (42.2) | 486 (47.3) |  |
| Rural | 853 (12.6) | 316 (13.5) | 166 (13.8) |  |
| **Insurance coverage (last 12 months)** |  |  |  | **<.0001** |
| No | 1959 (26.7) | 964 (35.3) | 349 (30.3) |  |
| Yes | 4693 (73.3) | 1515 (64.7) | 743 (69.7) |  |
| **Reproductive health wellness** |  |  |  | **<.0001** |
| No doctors visit | 2218 (32.6) | 848 (32.3) | 316 (28.7) |  |
| Doctor's visit (non-reproductive health) | 2997 (46.2) | 923 (40.3) | 435 (40.3) |  |
| Doctor's visit (reproductive health) | 1437 (21.2) | 708 (27.3) | 341 (31.0) |  |
| **Employment consistency** |  |  |  | **<.0001** |
| Not employed | 510 (6.1) | 199 (6.0) | 75 (6.3) |  |
| Employed (unstable) | 1342 (18.5) | 779 (30.7) | 245 (22.2) |  |
| Employed (stable) | 4800 (75.5) | 1501 (63.3) | 772 (71.5) |  |
| **PIR** |  |  |  | 0.9121 |
| Below 100% poverty | 1193 (15.3) | 463 (14.9) | 167 (14.7) |  |
| Above 100% poverty | 5459 (84.7) | 2016 (85.1) | 925 (85.3) |  |
| **Education status** |  |  |  | **<.0001** |
| Less than High School | 728 (10.8) | 266 (8.9) | 107 (8.4) |  |
| High School | 1959 (26.5) | 875 (32.4) | 374 (32.7) |  |
| Some college | 2079 (30.4) | 861 (36.0) | 380 (36.0) |  |
| College or more | 1886 (32.2) | 477 (22.7) | 231 (22.9) |  |

Table 3. Continued.
| Variables | Future fertility intentions |  |  | P-value |
| --- | --- | --- | --- | --- |
|  | Lowest risk<br>(n=6,652) | Sexual risk takers<br>(n=2,479) | Substance users<br>(n=1092) |  |
| **Race** |  |  |  | **<.0001** |
| NH White | 3455 (57.5) | 1313 (60.6) | 451 (47.4) |  |
| NH Black | 1121 (12.0) | 612 (16.5) | 364 (26.1) |  |
| Hispanic | 1533 (22.2) | 455 (17.6) | 233 (22.8) |  |
| Other | 543 (8.3) | 99 (5.3) | 44 (3.7) |  |
| **Immigration status** |  |  |  | **<.0001** |
| Non-immigrant | 5236 (77.9) | 2298 (92.2) | 933 (84.1) |  |
| Immigrant | 1416 (22.1) | 181 (7.8) | 159 (15.9) |  |
| **Union type** |  |  |  | **<.0001** |
| Married | 2640 (47.7) | 251 (12.2) | 28 (2.4) |  |
| Separated/ widowed/ divorced | 682 (7.4) | 241 (7.9) | 226 (18.7) |  |
| Never married | 3330 (44.9) | 1987 (79.9) | 838 (78.9) |  |
| **Number of biological kids** |  |  |  | **<.0001** |
| None | 3377 (48.1) | 1666 (71.2) | 680 (68.4) |  |
| At least 1 | 3275 (51.9) | 813 (28.8) | 412 (31.6) |  |
| **Age** |  |  |  | **<.0001** |
| Emerging adult (20-29 years) | 2689 (40.1) | 1465 (64.1) | 566 (58.7) |  |
| Adults (30 to 44 years) | 3963 (59.9) | 1014 (35.9) | 526 (41.3) |  |
| **Age at first sex** |  |  |  | **<.0001** |
| Early debut (< 15 years) | 1077 (14.3) | 683 (24.3) | 304 (23.7) |  |
| Non-early debut (>15 years) | 5575 (85.7) | 1796 (75.7) | 788 (76.3) |  |

MSA= Metropolitan Statistical Area; PIR= Poverty Income Ratio; NH=non-Hispanic

### Crude and adjusted multinomial regression models

Table 4 presents the result from the multinomial regression models fitted using the “lowest risk” level of the *PhP* variable as the outcome reference category to estimate effects of the independent variables on the “sexual risk takers” and “substance user’s” categories. All models were adjusted for the covariates: union type, number of biological children, participant age and age of sexual debut. The survey year was further included in the models to account for any temporal variations in the exposures and outcome over time.

**Table 4.** Social determinants of health and correlates of men’s preconception health status latent class membership (N=10,223).

|  | Crude model |  | Adjusted model |  |
| --- | --- | --- | --- | --- |
|  | Sexual Risk<br>Takers<br>vs<br>Lowest risk | Substance Users<br>vs<br>Lowest risk | Sexual Risk<br>Takers<br>vs<br>Lowest risk | Substance Users<br>vs<br>Lowest risk |
| **Metropolitan area** |  |  |  |  |
| Rural | 1 | 1 | 1 | 1 |
| Urban | 0.97<br>(0.72, 1.31) | 1.13<br>(0.88, 1.45) | 0.99<br>(0.71, 1.40) | 1.14<br>(0.89, 1.47) |
| Sub-urban | 0.85<br>(0.63, 1.14) | **0.77**<br>**(0.60, 0.99)** | 1.01<br>(0.72, 1.42) | 0.88<br>(0.68, 1.32) |
| **Insurance coverage**<br>**(last 12 months)** |  |  |  |  |
| Yes | 1 | 1 | 1 | 1 |
| No | 1.19<br>(0.97, 1.46) | **1.50**<br>**(1.27, 1.76)** | **1.25**<br>**(1.02, 1.52)** | **0.84**<br>**(0.71, 0.99)** |
| **Reproductive wellness** |  |  |  |  |
| Doctor's visit (reproductive health) | 1 | 1 | 1 | 1 |
| Doctor's visit (non-reproductive health) | **0.60**<br>**(0.47, 0.75)** | **0.68**<br>**(0.57, 0.81)** | **0.71**<br>**(0.55, 0.90)** | **0.82**<br>**(0.68, 0.98)** |
| No doctors visit | **0.60**<br>**(0.47, 0.77)** | **0.77**<br>**(0.64, 0.93)** | **0.59**<br>**(0.45, 0.76)** | **0.79**<br>**(0.65, 0.97)** |
| **Employment consistency** |  |  |  |  |
| Employed (stable) | 1 | 1 | 1 | 1 |
| Not employed | 1.11<br>(0.76, 1.61) | 1.19<br>(0.91, 1.55) | 0.68<br>(0.46, 1.00) | 0.83<br>(0.63, 1.10) |
| Employed (unstable) | **1.27**<br>**(1.04, 1.54)** | **1.98**<br>**(1.71, 2.30)** | **0.73**<br>**(0.59, 0.90)** | **1.23**<br>**(1.04, 1.45)** |
| **PIR** |  |  |  |  |
| Above 100% poverty | 1 | 1 | 1 | 1 |
| Below 100% poverty | 0.96<br>(0.74, 1.24) | 0.97<br>(0.81, 1.67) | **0.64**<br>**(0.49, 0.85)** | **0.73**<br>**(0.60, 0.89)** |
| **Education status** |  |  |  |  |
| Less than high school | 1 | 1 | 1 | 1 |
| High school | **1.59**<br>**(1.15, 2.20)** | **1.49**<br>**(1.17, 1.90)** | **1.74**<br>**(1.23, 2.48)** | **1.48**<br>**(1.15, 1.90)** |
| Some college | **1.53**<br>**(1.10, 2.14)** | **1.44**<br>**(1.12, 1.87)** | **1.65**<br>**(1.16, 2.36)** | **1.36**<br>**(1.05, 1.75)** |
| College or more | 0.92<br>(0.67, 1.26) | 0.86<br>(0.64, 1.15) | **1.94**<br>**(1.34, 2.79)** | **1.45**<br>**(1.09, 1.94)** |
| **Race/Ethnicity** |  |  |  |  |
| Non-Hispanic White | 1 | 1 | 1 | 1 |
| Non-Hispanic Black | **2.64** | **1.31** | **1.99** | 0.96 |
|  | **(2.12, 3.29)** | **(1.07, 1.60)** | **(1.55, 2.54)** | (0.79, 1.17) |
| Hispanic | 1.25 | **0.75** | 1.23 | **0.71** |
|  | (0.98, 1.59) | **(0.62, 0.91)** | (0.94, 1.59) | **(0.58, 0.87)** |
| Other | **0.54** | **0.61** | **0.62** | **0.70** |
|  | **(0.34, 0.86)** | **(0.43, 0.86)** | **(0.39, 0.99)** | **(0.51, 0.97)** |
| **Immigration status** |  |  |  |  |
| Non-immigrant | 1 | 1 | 1 | 1 |
| Immigrant | **0.67** | **0.30** | 0.90 | **0.39** |
|  | **(0.52, 0.85)** | **(0.23, 0.39)** | (0.68, 1.18) | **(0.29, 0.52)** |

Health access, socioeconomic status and sociocultural contexts but not residential status were significant predictors of class membership. Men’s access to health insurance coverage and type of health care service received were significant predictors of men’s preconception health status. A lack of health insurance coverage was associated with 16% reduced odds for belonging to the “substance users” (aOR:0.84, 95%CI:0.71, 0.99) and 25% increased odds for belonging in the “sexual risk takers” category than in the “lowest risk” category compared to those who had insurance coverage (aOR: 1.49, 95%CI: 1.19, 1.86). Whereas not having a doctor’s visit or having a non-reproductive health visit were associated with reduced odds for belonging either the sexual risk takers or substance users’ categories than the lowest risk class

Employment consistency, poverty level and educational status were associated with men’s preconception health status. Employment instability was associated with reduced odds for belonging in the “sexual risk takers” category (aOR: 0.73, 95% CI: 0.59, 0.90) than to the “lowest risk category but increased odds for belonging in the “substance users” category (aOR: 1.23: 95% CI: 1.04, 1.45) than in the “lowest risk” category. Unemployed status did not have a statistically significant effect on preconception health class membership. Living below 100% poverty level reduced the odds for belonging in the “sexual risk takers” and “substance users” categories by 36% and 27% respectively (aOR: 0.64, 95%CI 0.49, 0.85; aOR: 0.73, 95% CI: 0.60, 0.89 respectively). Education status was significantly associated with men’s preconception health status. Compared to those with less than high school education, having a high school degree, some college or a college degree or more was associated with increased odds of belonging in the “sexual risk takers” category (aOR: 1.74, 95% CI: 1.23, 2.48; aOR: 1.65, 95% CI:1.16, 2.36; aOR: 1.94, 95% CI1.34, 2.79 respectively) and the “substance users” category (aOR:1.48, 95% CI: 1.15, 1.90; aOR:1.36, 95% CI:1.05, 1.75 and aOR:1.45, 95% CI:1.09, 1.94 respectively).

Racial and ethnic background was also a significant predictor of preconception health status latent class belonging. Compared to their non-Hispanic White counterparts, being non-Hispanic Black was associated with 99% increased odds of belonging in the “sexual risk takers” category (aOR:1.99, 95% CI: 1.55, 2.54) than to the “lowest risk” category. Whereas, identifying as Hispanic was associated with 29% reduced odds of belonging in the “substance users” category (aOR:0.71; 95%CI: 0.58, 0.87) than in the “lowest risk” category. Immigration status was also a significant determinant of men’s preconception health status. Compared to non-immigrants, immigrant status was associated with reduced odds for belonging in the “substance users” category (aOR=0.39; 95%CI: 0.29, 0.52) than in the “lowest risk category.

## DISCUSSION

This population-level study utilized pooled cross-sectional data from the 2011-2019 NSFG dataset to identify latent classes of men’s preconception health and examine associations between these latent classes and the social determinants of health – health access, socioeconomic status, and social and community factors. Per the hypothesis of this study, three latent classes of men’s preconception health were identified – low-risk, substance users and sexual risk taker categories and these classes were predicted by all the social determinants of health domains except for men’s residential context. The salience of these findings are discussed in the following paragraphs

This study found that 22% of all men belonged to the “substance users” class and almost 10% belonged in the “sexual risk-takers.” The latent classes observed in this study, specifically on substance use and risky sexual behaviors, may not be a function of “free choice” but a constellation of the individual, structural, contextual, and social factors, including hegemonic practices, that have intertwined over the life course to define masculinity and masculine behaviors. There is a gendered narrative to risk-taking and risk behaviors with men more likely to engage in risky sexual behaviors or substance use than women [35]. Our study supports this narrative to the extent that distinct categories of men within both risk categories were explicitly identified. This finding, however, continues to raise questions about the structures that have created and continue to perpetuate masculinity as inherently risky. Given the risks for adverse health outcomes for men, due to substance use and risky sexual behaviors, its potential influence on their partners health and wellbeing, and the well documented negative outcomes for pregnancies and families [5, 15, 36], concerted efforts are needed to better engage men in preconceptional health care more holistically.

Rural/urban residence was not associated with men’s preconception health status. This finding was contrary to a seminal study that used the NSFG data and found men’s preconception health status to differ by rural/urban residence [26]. This difference may be explained by Choiiyyah and colleagues’ focus on men who needed preconception care (i.e., men intending a pregnancy). The current study provides additional insight by examining all fecund men between 20 to 44 years, independent of their fertility intentions. Because about 40% of men report unintended pregnancies [37], this approach allowed the examination of risk exposure and background differences independent of pregnancy intentions. Rural/urban residence alone has well acknowledged limitations in examining the nuances of residential context and its contributions to poorer health. Among men specifically, Thorpe et al. (2015) argued for a place-based approach that examines the social environment in which men live to create a better understanding of the effect of place on men’s health in general and on their preconception health specifically [38]. More studies are needed that examine the social contexts, particularly neighborhood factors – such as social cohesion and racial composition - in which men live and plan to establish families.

Although most men in this study had health insurance, the majority, across all latent classes did not have a reproductive health visit. This finding is consistent with previous studies in which men were found to have increased access to health care through insurance coverage, yet did not necessarily translate into increased access to reproductive health services [12]. It is plausible that men may be uncomfortable with discussing their reproductive health or see it as an inherently female domain, hence they are less likely to utilize these services [39]. Another thought could be that health care providers are implicitly biased towards providing reproductive health services for females rather than for males [40, 41]. It is plausible that providers’ bias towards men’s reproductive health screening may make them less inclined to inquire about men’s reproductive health if they (men) did not exhibit or report sexual risk taking explicitly. It is also plausible that, men in high-risk preconception health categories may be more inclined to seek reproductive care compared to other men in lower risk categories which may explain the reduced odds of non-reproductive health utilization. Given the preconception health phenotypes of men in this study, identifying the barriers men face to seeking care, beyond access to health insurance, and finding ways to surmount these barriers should remain a public health priority. Several studies and reports have emphasized the value of effectively incorporating reproductive health screening into men’s routine health care visits [12, 42, 43]. By making intentional changes towards ensuring that each health care encounter, specifically with men between ages 20 to 44 years, provides an opportunity for reproductive wellness, practitioners can begin to improve men’s health status preconceptionally.

This study found that men living below the poverty line were less likely to belong to the “sexual risk takers” or “substance users” categories which contradicts previous findings. Previous studies showed that men who have a higher education status were more likely to make health behavior changes preconceptionally [44]. Specifically, Marcell et al. (2016) did not find any association between poverty status and men’s need for family planning [14]. It is often thought that increased socioeconomic status can reduce stress for men and can be motivational towards starting a family [5]. However, in some contexts, financial independence increases the chance for risky behaviors due to perceptions of what masculine behaviors should look like for those in higher income brackets [45]. Substance use and sexual risk taking may be more difficult to afford at the lower ends of socioeconomic status. Conversely, men with a high school degree and higher were more likely to belong in the higher risk latent classes. Previous studies examining men’s preconception health suggest that men with higher education status are more likely to make health behavior changes preconceptionally [44]. One explanation of this finding could be stressors related to achievement and career advancement which may lead men to engage in more risky behaviors as a coping strategy [45]. An alternative explanation could be the accumulation of wealth and negative masculine perceptions which may influence men to engage in risky behaviors as a sign of manhood or masculinity [35]. More studies are needed to examine the contexts of men’s masculinities and their influence on men’s health behaviors. Similar findings were identified with employment status which suggests that changes in employment can influence the social class and masculinities pathway. Men often perceive themselves or have traditionally been socially constructed as “breadwinners” [46]. Unstable employment can impact men’s perceptions of themselves as breadwinners, consequently increasing the likelihood of engaging in risky behaviors, including substance use[45].

Men’s immigration status was associated with decreased odds for belonging in any of the high-risk preconception health phenotypes. This finding was expected given that many other studies that explored immigration status on health often indicate the “healthy immigrant” paradox, in which immigrants to the US are healthier overall than the general US population [47]. An unexpected finding, however, was the increased odds for belonging to the sexual risk takers category than the substance user’s category among immigrants. Since this study examined all immigrants, it limits our understanding of immigrants’ preconception health statuses at the intersection of race/ethnic belonging. Further, given the context of acculturation, through which immigrants are thought to lose their health advantage over time, there is need for more studies to explore – alongside racial/ethnic backgrounds – how acculturation in male populations impacts their preconception health. Future studies will benefit from approaches that examine how long immigrants have lived in the US and variations in preconception health outcomes by racial/ethnic belonging.

### Limitations and Implications for Future Research

This study was limited by several factors. First, it utilized cross sectional data which limits any causal associations between the exposure and outcome. Second, only a few preconception risk factors were examined. For instance, measures on smoking, high blood pressure and other preconception health indicators were not utilized because they were either not measured in the dataset, or were inconsistently measured across survey years. While the NSFG data was well suited to answer the questions posed by this study, preconception indicators could be grossly understated. Further, all variables utilized in this study were self-reported, which can introduce social desirability, reporting and recall biases.

There were also limitations to this study in terms of the social determinants of health measures that were utilized. For instance, immigration status was measured as a binary variable (i.e., whether or not a man was an immigrant or not) which is less informative than the length of time an immigrant had spent in the U.S. The length of time an immigrant spends in a host country has been associated with acculturation and stresses which can, in turn, impact preconception health. Future studies should consider using earlier years of NSFG data to explore these phenomena. This study provides valuable insight to men’s preconception health; however, the data source does not capture measures of masculinity which limits our understanding of why we observed these results. Future studies will benefit from qualitatively examining these phenomena among men, specifically exploring the factors that contribute to reduced contraceptive use, perceptions of health, and its influence on fatherhood.

## CONCLUSION

Taken together, men’s health access, socioeconomic status and sociocultural contexts can collectively have meaningful impacts on their preconception health, even though it may be through different pathways. In some contexts, manliness is expressed through men’s ability to engage in risky behaviors, including substance use and sexual promiscuity, while also remaining financially successful as demonstrated by achievement in educational advancement and social mobility. This study underscores the need to explore more in-depth the relationship between social class, masculinity, and men’s preconception health. This study suggests that other underlying factors that are nested in men’s experiences of their social contexts influence men’s preconception health, however nationally representative datasets are not designed to measure these unique experiences. Studies that examine how men, on the cusp of fatherhood, view themselves, their roles as fathers and their health statuses at this intersection can improve our knowledge of the socio-cultural processes that intertwine to shape men’s preconception health.

## Data Availability

The data used in this study are publicly available through the National Center for Health Statistics website. The datasets generated and/or analyzed during the current study are available in the Harvard Dataverse repository, https://doi.org/10.7910/DVN/4X4IEG.

