## Supplemental Tables 1a and b for "Men’s preconception health and the social determinants of health: What are we missing?"

**Supplementary material**

eTable 1a. Survey weighted relative measures of fit for men's preconception health phenotypes.

| **Model fit Indices** |  |  |  |  |  |
| --- | --- | --- | --- | --- | --- |
| No. of latent classes | 2Log-Likelihood | G-squared | BIC | Entropy | Change in BIC |
| 2 class model | -33076.35 | 577.18 | 734.13 | 0.48 | 1,458.14 |
| **3-class model** | **-33015.12** | **454.72** | **694.76** | **0.82** | **39.37** |
| 4-class model | -32962.24 | 348.96 | 672.1 | 0.44 | 22.66 |
| 5-class model | -32924.67 | 273.81 | 680.03 | 0.66 | -7.93 |

eTable 1b. Estimation of mean membership probability and class size, 3-Class model.

| Latent classes | Class size | LC1 | LC2 | LC3 |
| --- | --- | --- | --- | --- |
| LC1 | 1092 | **0.972** | 0.007 | 0.009 |
| LC2 | 6652 | 0.009 | **0.931** | 0.145 |
| LC3 | 2479 | 0.018 | 0.062 | **0.845** |

*LC: Latent class*
